# It’s not the shimmy, it’s the shift: Differential effects of valence shift type and stimulation mode during a simulated EMDR session in PTSD patients and healthy controls

**DOI:** 10.64898/2026.01.08.26343688

**Authors:** Valeska Pape, Francine Barczyk, Caroline von Klitzing, Carla Fitting, Markus Stingl, Eva Schäflein, Olaf Wolkenhauer

## Abstract

**Introduction:** Mechanism-of-action studies on Eye Movement Desensitization and Reprocessing have so far focused mainly on the presumed active component of bilateral stimulation (BLS). In this pilot study, a further potential working mechanism was examined for the first time, involving stimulation-induced changes in emotional valence.

**Methods:** Twenty-five patients with posttraumatic stress disorder and 25 healthy controls between 19 and 64 years of age underwent a simulated intervention based on components of Eye Movement Desensitization and Reprocessing (EMDR). Each participant was presented with 18 individual script pairs, simulating different valence shifts (valence switch into neutral, valence switch into positive, no valence shift), whereas BLS vs. no BLS were applied. During the intervention, subjective and physiological emotional responses were measured.

**Results:** When valence shifted to positive or neutral, a significant change in treatment-relevant subjective and physiological effect measures was found compared to scripts without a valence shift. For stimulation type, no subjective, but significant physiological effects were observed: The controls showed a physiological de-arousal under BLS, indicated by a decreased skin conductance level, and the patients showed an accelerated heart rate and an increased M. zygomaticus activity. Significant interaction effects were observed: Under BLS, the arousal-reducing and valence-changing effects of negative to neutral switches increased. Interestingly, these BLS effects became conscious to the participants only when valence switches were applied.

**Discussion:** The findings provide new insights into the potential emotion-modulating physiological effects of BLS and its interplay with changes in emotional valence.

## INTRODUCTION

In 1989, a new, gentle method for the treatment of posttraumatic stress disorder (PTSD) was developed (Boudewyns and Hyer, 1996; Chen et al., 2014; Shapiro, 1989; Shapiro, 2014; Shapiro, 2017; Valiente-Gómez et al., 2017), known as Eye Movement Desensitization and Reprocessing (EMDR). The therapy is well established, empirically validated (Cuijpers et al., 2020; Van Etten and Taylor, 1998; Jericho, Luo, and Berle, 2022; Khan et al., 2018; Lewey et al., 2018; Mavranezouli et al., 2020) and applicable for the first-line treatment of PTSD and a broad range of further psychiatric disorders (Hase, 2006; Hase, Schallmayer, and Sack, 2008; Hofmann, 2014; Knipe, 2008; Stingl et al, 2022).

### EMDR standard protocol

The procedure of a typical session is described in Shapiro’s standard protocol (Shapiro, 2017). The confrontational phase starts with a brief activation of the worst moment of the memory and an initial rating on the Subjective Units of Distress Scale (SUD, from 0 = minimally distressing to 10 = maximally disturbing) and the Validity of Cognition Scale (VoC, from 1 = minimal credibility to 7 = maximum credibility of an event-related positive cognition [PC]). Thereafter, the patient is asked to silently follow her spontaneous associations while bilateral stimulation (BLS) is applied. After a certain number of BLS, the therapist interrupts the chain of associations with the question, ‘What happened last?’. The further procedure depends on the emotional valence of this previous association: If it is a negatively connoted association (*no change in valence*), another set of BLS is performed. If it is a neutral or positively connoted association twice in a row (*change in valence)*, the stimulation is stopped, and the association path is considered complete. Ultimately, the therapist returns to the traumatic event, and the SUD and VoC ratings are collected again. If the SUD value decreases sufficiently (e.g., to 1 or less) and the VoC value increases sufficiently (e.g., to 6 or higher), the PC is reinforced with slow BLS, and the session is considered completed. If the decrease or increase is insufficient, further fast BLS is used until the target criteria are met. Otherwise, the session is rated as ‘incomplete,’ and the trauma topic is revisited in the next session.

### Mechanism of action

Since its inception, EMDR has been the subject of intensive research. Based on a growing number of experimental studies, various models of action have been developed. The working memory model (Andrade, Kavanagh, and Baddeley, 1997; Wadji, Martin-Soelch, and Camos, 2022) as well as physiological explanations (Armstrong and Vaughan, 1996) have proven to be the most valid (Landin-Romero et al., 2018).

The *working memory model* (Andrade et al., 1997; Baddeley & Hitch, 1974) explains the therapeutic effect of EMDR through a dual attention process: If attention is simultaneously diverted to two concurring tasks – such as BLS (task 1) while imagining a traumatic situation (task 2) -, the limited capacity of working memory may be insufficient to experience the emotions in all its severity, leading to a subjective reduction in stress (Haymann, 2008). This effect has been demonstrated in various studies (Houben et al., 2020; Maxfield, Melnyk, and Haymann, et al., 2008; Mertens, Lund, & Engelhard, 2020; Servan-Schreiber et al., 2006), with the dual attention effect being evident not only in BLS tasks but also in other tasks – even those not used within EMDR (Gunter and Bodner, 2008; Engelhard, van den Hout, and Smeets, 2011; van den Hout et al., 2011; Tadmor, McNally, & Engelhard, 2016; Engelhard, Van Uijen et al., 2010).

*Psychophysiological models*, in turn, explain the therapeutic effect of EMDR through a BLS-induced intensified orienting response (Armstrong & Vaughan, 1996; Sokolov, 2001), which is defined as an involuntary increase in attention following new or changing stimuli. It is controlled by brain areas, such as the superior colliculus (Benedetti, 1995). These effects can be measured by parameters of sympathetic activity (e.g., skin conductance response [SCR] and heart rate [HR]). If sensory stimuli are applied repetitively, an intensified orienting response may occur, followed by a parasympathetic relaxation or habituation response resulting from the loss of novelty of the stimuli. There are diverse studies demonstrating such an initial increase in SCR and HR under BLS in PTSD patients, followed by relaxation (Wilson, Silver, Covi, & Foster, 1996; Sack et al., 2008). A direct SCL- and HR-decrease under BLS in PTSD patients was observed as well (Elofsson, von Scheele, Theorell, & Söndergaard, 2008; Montgomery & Ayllon, 1994; Sack et al., 2008; Schubert et al., 2010). Barrowcliff et al. (2004) observed comparable effects in healthy individuals, and Jauch et al. (2023) demonstrated inhibited amygdala activity under BLS in mice. In a study by Reichel et al. (2021), a lower startle activity for negative standard scripts, an increased SCR activity for positive scripts, and a lower general startle sensitivity were found under BLS compared to monolateral stimulation and no BLS. These findings, however, were restricted to healthy individuals and could not be replicated in PTSD patients (Pape et al., 2024).

### Simple vs. extended working model

What all these component studies have in common is their restriction to the presumed effective factor of BLS (*simple effective model*, Fig. 1). In her Adaptive Information Processing Model (AIP), Shapiro (2006, 2007) emphasized that the necessary condition for trauma-focused reprocessing is thus not the BLS per se, but rather a BLS-induced adaptive association process (i.e. associating pathogenic memories with memory networks containing adaptive information) that induces a valence shift (*extended effective model*, Fig. 1). This valence shift can be observed on various levels (cognitions, emotions, bodily sensations), but always involves a change from initially negative and self-destructive aspects to neutral or positive ones. Table 1 illustrates examples of valence shifts on these different levels, contrasted with no valence shift, i.e., a series of persistent negative associations (in EMDR known as ‘cycling’). This approach is not a novel one, but can also be found in other treatments such as trauma-focused Cognitive Behavior Therapy (tf-CBT), for example, in the context of the ABC model (Ellis, 1991) and the technique called cognitive reprocessing (Wilken, 1998). Still, the connection of valence shifts with BLS as a ‘shifting motor’ is novel and innovative. So far, however, there are no studies that examine the effects of BLS on valence-shifting processes in patients with PTSD, and the exact and reciprocal influence of both factors on the intervention success remains unclear.

**Figure 1:**
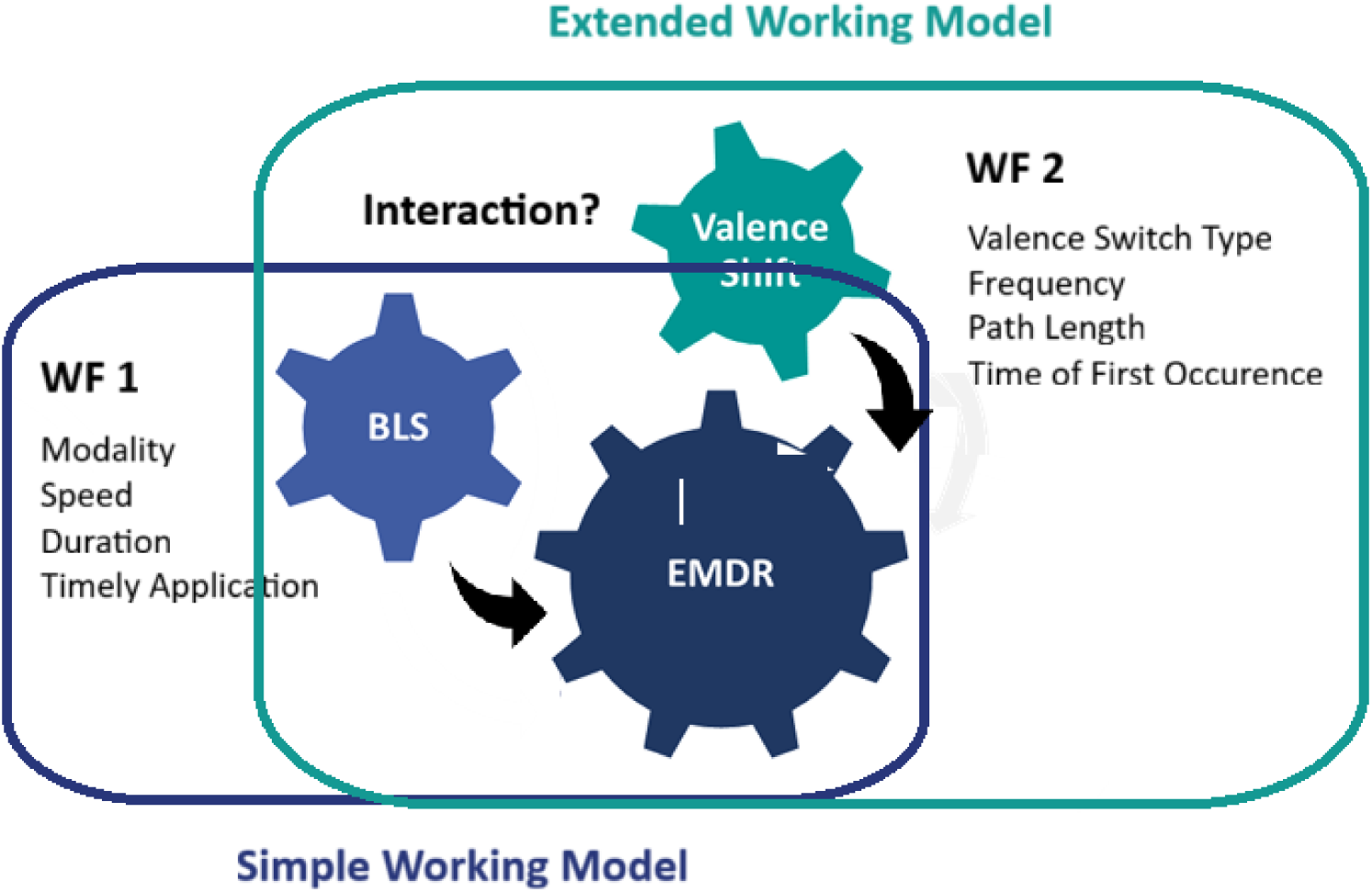
Simple vs. extended working model

**Table 1:**
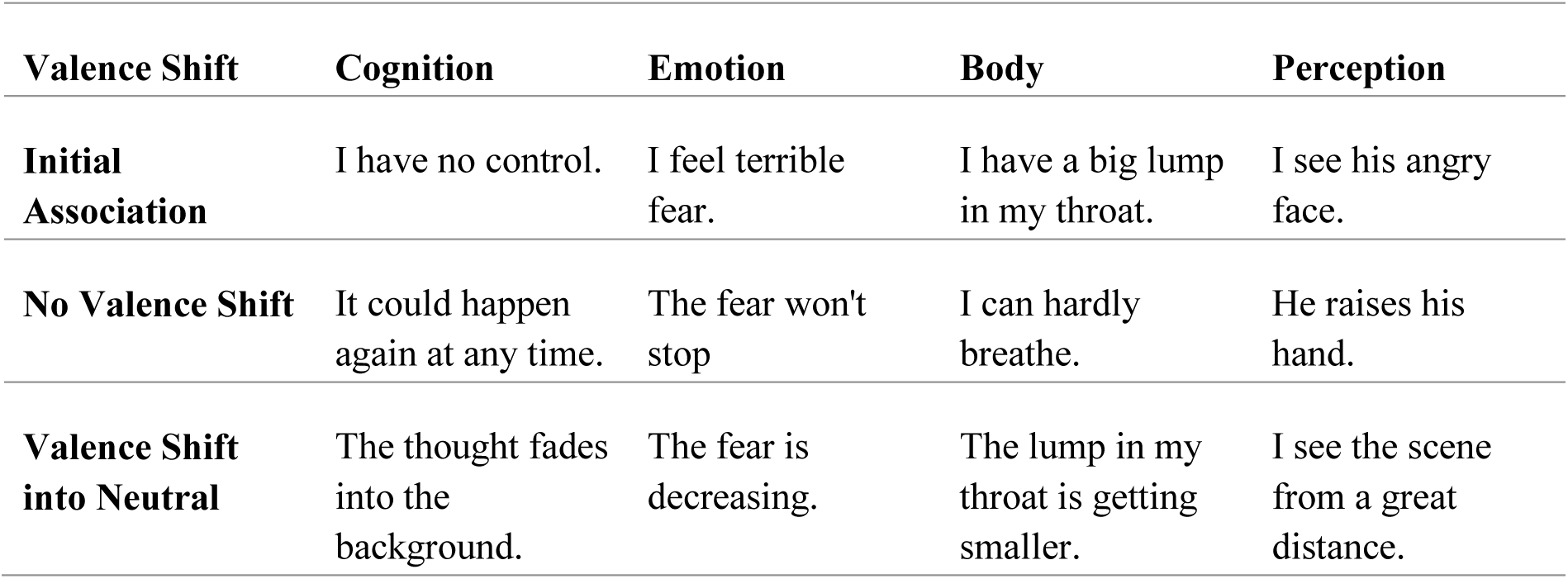
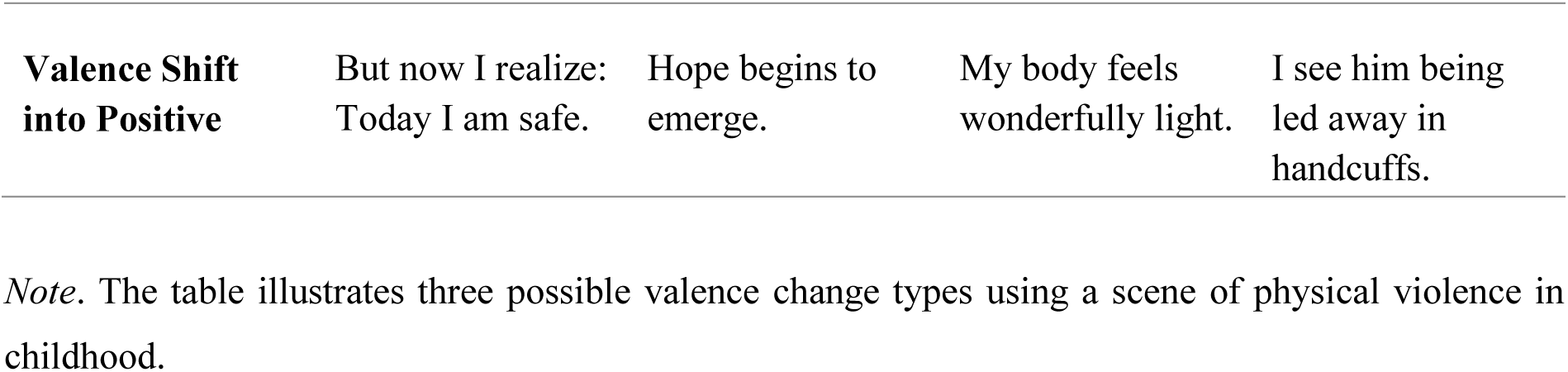
Valence shift types.

| Valence Shift | Cognition | Emotion | Body | Perception |
| --- | --- | --- | --- | --- |
| **Initial Association** | I have no control. | I feel terrible fear. | I have a big lump in my throat. | I see his angry face. |
| **No Valence Shift** | It could happen again at any time. | The fear won't stop | I can hardly breathe. | He raises his hand. |
| **Valence Shift into Neutral** | The thought fades into the background. | The fear is decreasing. | The lump in my throat is getting smaller. | I see the scene from a great distance. |
| **Valence Shift**<br>**into Positive** | But now I realize:<br>Today I am safe. | Hope begins to<br>emerge. | My body feels<br>wonderfully light. | I see him being<br>led away in<br>handcuffs. |

*Note.* The table illustrates three possible valence change types using a scene of physical violence in childhood.

## AIMS AND HYPOTHESES

### Aims

This experimental pilot study aims to examine the influence of stimulation type (BLS vs. no BLS), valence switch type (switches into positive, switches into neutral, no valence switch), and interactions of both on the effect of a simulated EMDR session for the first time. This will be done by collecting a data set of 25 PTSD patients and 25 healthy control subjects. They will undergo a simulated session with a fixed variation of different valence shifts and stimulation types. Subjective and physiological stress, as well as resource parameters, will be used to measure the success of the intervention.

### Hypotheses

H1: The stimulation type (BLS vs. non BLS) influences intervention success (subjectively*, physiologically**).

H2: The valence shift type (valence shift vs. no valence shift) influences intervention success (subjectively*, physiologically**).

H3: BLS and valence shift type mutually influence each other (stronger subjective* and physiological** valence shift effects under BLS compared to no BLS).

* subjective: decrease in the SUD score (less subjective distress) and the SAM arousal score (less subjective arousal), increase in the VoC score (i.e. a stronger validity of the PC) and the SAM valence score (more positive feelings)

** physiological: decrease in the Skin Conductance Level (SCL), HR (physiological de-arousal), and M. corrugator activity (less frowning), increase in M. zygomaticus activity (more smiling)

### Sample

25 PTSD patients and 25 healthy controls without psychiatric disorders between 19 and 64 years of age were included in the study. Both groups were matched for age, sex, and education level. All participants gave their written informed consent. The demographic and clinical characteristics of the sample are depicted in Table 2.

**Table 2:** Demographic and clinical sample characteristics

|  | Overall | Patients | Controls | PPG vs. CG |
| --- | --- | --- | --- | --- |
| **Age (years)** | 33.35 (13.19) | 31.52 (11.15) | 34.96 (14.80) | .368 |
| **Sex** | 34 females<br>16 males | 19 females<br>6 males | 15 females<br>10 males | .353 |
| **Education** | 26 high-level<br>21 middle level<br>3 low-level | 10 high-level<br>12 middle level<br>3 low-level | 16 high-level<br>9 middle level<br>0 low level | .196 |
| **VAS Mood** | 4.26 (1.78) | 5.00 (1.54) | 3.54 (1.72) | .004** |
| **VAS Arousal** | 4.83 (2.27) | 5.35 (2.10) | 4.33 (2.35) | .127 |
| **IES** | 0.30 (0.46) | 0.57 (0.51) | 0.06 (0.23) | < .001*** |

*Note.* IES = Impact of Event Scale. Values are means, standard deviations, or numbers. Group differences in age and the screening scales are tested via univariate ANOVAs (two-tailed). Group differences in gender and education level are tested using a chi-squared test (two-tailed). \*\* $p < 0.01$ ; \*\*\* $p < 0.001$ .

#### PTSD patients

The patients were recruited from inpatient and outpatient therapy programs of the Department of Psychiatry and Psychotherapy, University Medicine Rostock (UMR). The inclusion criterion was the presence of a posttraumatic stress disorder (ICD-10: F43.1) as the main diagnosis. The mean time since the stressful event was 20.77 months (± 7.50 months). There were 17 patients with a history of poly-traumatization (Type 2 trauma) and six patients with mono-traumatization (Type 1 trauma). 21 patients showed clinically relevant depressive symptoms (BDI > 13), which was accepted due to the well-known high overlap of depressive and PTSD symptoms (Gros et al., 2012). Eight patients had already participated in trauma therapy (one patient in EMDR). Nine patients were inpatients, six patients were outpatients, and ten had no current treatment at the time of the study. Comorbid disorders such as mild to moderate depression, anxiety disorders, and substance use disorders were deliberately included in the study. Ethical approval was obtained from the ethical committee of the University of Medicine Rostock.

#### Control subjects

Recruitment methods for the control subjects included advertisements in newspapers, newsletters, and postings on the university campus. The inclusion criterion was the absence of any psychiatric diagnosis described in the ICD-10.

#### Exclusion criteria

PTSD criteria were assessed using the SKID I (Structured Clinical Inventory of Psychiatric Disorders for DSM-IV; Wittchen, Zaudig, and Fydrich, 1997). In addition, the Impact of Event Scale (IES; Weiß and Marmar, 1997) was used. Individuals with documented severe mental disorders (drug abuse, severe depression, dementia, schizophrenic psychosis, and intellectual disability retardation), neurological diseases (such as seizures in the anamnesis), severe hearing or visual disabilities, medication with influence on startle reflex response (benzodiazepines, buspirone, opioids), recent medication switchover within the last 2 weeks, or insufficient knowledge of German were excluded from the study. All psychiatric diagnoses were based on ICD-10 criteria and established by experienced clinical raters.

### Procedure

***Preparation (60 min).*** After being informed about the purpose and procedure of the study and providing written and verbal consent, participants completed the screening questionnaires and reviewed the inclusion and exclusion criteria.

***Interview (60 min):*** At the beginning, a stressful event (SUD score > 6) was selected for the simulated session. Further details were obtained in a semi-structured interview, which was needed to create the individual scripts. The worst moment of the stressful event was determined, and associated images, thoughts, feelings, bodily sensations, and positive counter-associations were elicited (e.g., ‘the perpetrator’s aftershave’ vs. ‘vanilla smell’, ‘I have no control’ vs. ‘Today I am safe’). At the end of this part, the participant also chose the event-related PK for the simulated session.

***Script creation (by the experimenter):*** The experimenter subsequently created 36 individual scripts, sorted into 18 script pairs. For details see the section ‘Stimulus material.’

***Simulated session (60 min):*** The simulated EMDR session was conducted as a stimulus-response paradigm without the presence of a therapist. The setting was experimental, allowing for different types of valence shifts (no valence shift, valence shift to neutral, and valence shift to positive) and stimulation types (with and without BLS) to be simulated while keeping other factors constant. This allowed us to specifically investigate the influence of both variables on intervention outcome. The 3 x 2 intervention conditions (valence shift x stimulation type) were presented in six different orders to prevent order effects (Fig. 2).

**Figure 2:**
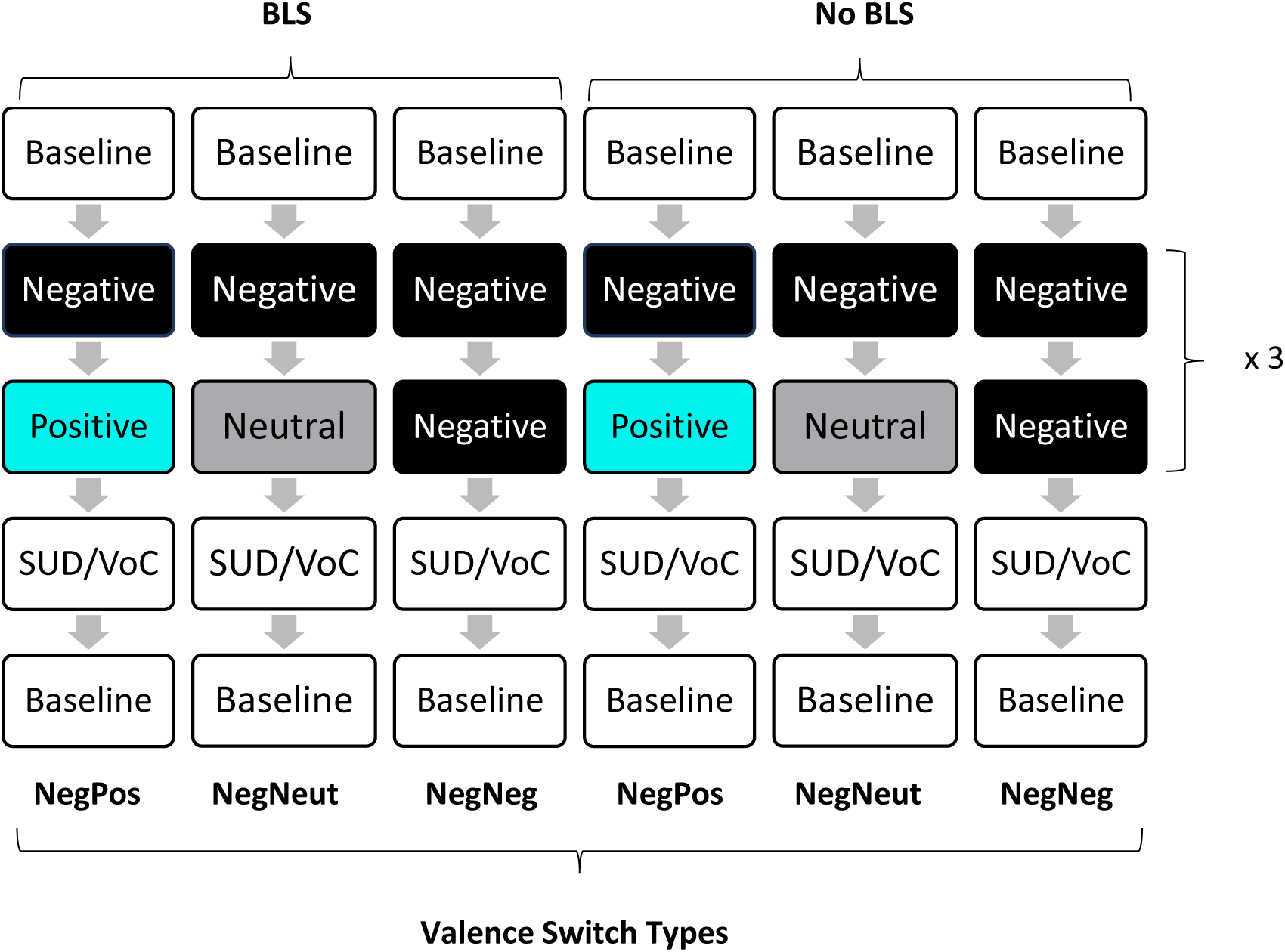
Procedure of the simulated session *Note.* The figure depicts the six intervention conditions. A series of three script pairs, each representing a valence switch/stimulation type category, was defined as a block. Each participant was confronted with all intervention conditions (within-group-design). Stimulation Types: BLS vs. No BLS. Valence Switch Types: NegPos (valence switch into positive), NegNeut (valence switch into neutral), NegNeg (no valence switch).

The participant was equipped with surface electrodes and was given two vibration pads (Deluxe Tac/Audioscan Device Revision 5.1 from NeuroTek Corporation) to trigger the tactile BLS. Subsequently, the 18 script pairs were presented in randomized order, interrupted by white screens, with half of them containing BLS and the other half without (Table 3). The presentation was conducted in a blocked format, with three script pairs of the same valence and stimulation type combined into a single block. A baseline measurement with a white screen (*Baseline*, 16 seconds) was taken between each block. The participant was tasked with reading the script contents (*Reading Task*, 6 seconds) and imagining them as accurately as possible (*Imagination Task*, 10 seconds). To determine the effect of each valence change × stimulation type intervention, the participant was asked to evaluate the current SUD and VoC value at the end of each block (*Rating Task*, 18 seconds). In the 7.5 seconds before and 10 seconds after the script transition, physiological parameters were recorded.

***Subjective Rating.*** At the end of the session, the 18 script pairs were presented again. To ensure that the scripts reflected the actual event-related associations of the participants, the consistency of the scripts was assessed using SAM-analog consistency scales. To ensure that scripts categorized as positive, neutral, or negative were indeed experienced in this way, the valence and arousal of each script were recorded using the scales of the SAM at the end of the session.

### Stimulus material

The affective stimulus material was developed based on a 60-minute interview. 36 individual scripts were created, sorted into 18 script pairs. The procedure was based on Pape et al. (2024), which is an adaptation of the method from Bichescu-Burian et al. (2018) for PTSD patients. Each single script consisted of 15 to 20 words and corresponded to one of three association types: negative, neutral, or positive. For all scripts, a constant reading- (6 seconds) and imagining time (10 seconds) was maintained. Via pairing, three different valence shift categories were simulated: NegPos (6 script pairs), NegNeut (6 script pairs), and NegNeg (6 script pairs). The first script in each pair was always negative (e.g., ‘I hear his gasping breath and see his greedy gaze’). In the second script, a worsening (e.g., ‘His lascivious face is getting closer and closer’), a neutralization (e.g., ‘His face is getting blurrier and blurrier until I can no longer recognize it’), or a change to a positive association (e.g., ‘The image changes until I see my grandmother’s lovely face in front of me’) were described. Within each valence shift category, two script pairs referred to emotions, two ones referred to thoughts, and two ones referred to bodily sensations.

### Stimulation mode

Stimulation was conducted tactilely using the Deluxe Tac/Audioscan Device, Revision 5.1, from NeuroTek Corporation, to avoid confusion with the EMG measurement. Rhythmically changing vibration signals were applied to the person’s palms (Contact NeuroTek Corporation, Wheat Ridge, CO). This instrument enabled the researcher to minimize experimenter effects by choosing a fixed duration and frequency of the vibration signals. Stimulation began at the start of each BLS block and continued until the end of the block. Two stimulation modes were used: BLS (where the right and left hands were bilaterally stimulated in fast alternation) and no BLS. To record changes in physiological baseline parameters, two-thirds of the white screens were also stimulated.

### Subjective measures

#### SUD Scale and VoC Scale

At the end of each block, current distress was assessed using the SUD-scale, which ranges from 0 (minimal stress) to 10 (maximum stress). In addition, the validity of the initially chosen PC was assessed using the VoC-scale, which ranges from 1 (minimal validity) to 7 (maximal validity). Both scales are part of the EMDR standard protocol (Shapiro, 2017) and are used here to determine the effect of each block, respectively, of each valence shift and stimulation type condition.

#### SAM

At the end of the entire session, subjective emotional responses to the script category conditions were rated on nine-point scales based on the SAM of Bradley and Lang (1994). The participants were instructed to reflect on and recall their feelings during the imagination task. Valence (ranging from 1 to 9, with negative to positive) and arousal (ranging from 1 to 9, with low to high arousal) of the emotional reactions were measured. To determine the consistency of the scripts with the actual memories, a SAM-analogue consistency scale (ranging from 1 to 9, with low to high consistency) was used.

#### Visual Analogue Scales (VAS)

Before the experiment, current mood (ranging from 1 to 100, with positive to negative mood) and arousal (ranging from 1 to 100, with low to high arousal) were measured via VAS (Gift, 1989).

### Physiological measures

#### Mimic muscle activity

Mimic muscle activity was measured as electromyogram (EMG) of the left corrugator and zygomaticus muscle using Ag/AgCl miniature electrodes following the recommendations by Fridlund and Cacioppo (1986). The MP160 Research System registered the raw EMG signal, filtered it with a 28–500 Hz Bandpass Filter, and then multiplied the signal by a factor of 100. The integrated EMG signal was digitally evaluated using AcqKnowledge Software from Biopac Systems. Baseline-corrected mimic muscle activity was defined as the difference between mean EMG (mean EMG value within 4 seconds after script onset) and baseline EMG (EMG value within the 0,1 second before script onset). The sampling rate was 1000 Hz. Signal quality was visually inspected throughout the recording session. Trials with excessive baseline activity, noise, or movement artefacts were excluded from the analysis.

#### Electrodermal activity (EDA)

EDA was derived via Ag/AgCl standard electrodes on the hypothenar muscle of the non-dominant hand. The signal was registered by the MP160 Research System and filtered with a 28–500 Hz Bandpass Filter. Digital input to stimulus events was extracted from all digital lines. Baseline-corrected ‘SCL’ was defined as the difference between mean SCL between the mean SCL (the mean SCL value within 900 to 4000 milliseconds after script onset) and the baseline SCL (the SCL value within the 1 second preceding script onset).

#### Electrocardiographic activity (ECG)

ECG was measured using Einthoven lead II, with Ag/AgCl standard electrodes. Raw analogue ECG signal was sampled at 1000 Hz and filtered through a 0.05–150 Hz band-pass filter. Thereafter, times of R peaks < 1 mV within the analogue signal were detected. According to the recommendations of Graham (1978), second-to-second HRs in beats per minute (bpm) for the 4 s after picture onset were computed. Three values were extracted: ‘overall HR change’ was defined as the difference between mean HR (mean second-by-second HR for the 4-s window after script onset) and baseline (mean HR for the 1 s before script onset), ‘HR acceleration’ was defined as the difference between HR_max_ (maximal second-by-second HR for the 4 s after script onset) and baseline, and ‘HR deceleration’ was defined as the difference between baseline and HR_min_ (minimal second-by-second HR for the 4 s after script onset). For trials in which HR never increased above baseline, the acceleration score was set to zero. Deceleration scores were handled analogously.

### Statistics

The data were statistically processed using IBM SPSS Statistics 22.0 Software. The raw data of each subject were averaged per intervention condition. The standard distribution requirement was checked visually. Comparisons of descriptive data and VAS values were performed using univariate analyses of variance (ANOVA) (two-tailed) or Chi²-tests.

A repeated-measures ANOVA was used to examine the subjective and physiological effects of each valence switch and stimulation type condition. ‘Stimulation type’ (BLS vs. No BLS), and ‘valence switch type’ (NegPos, NegNeut, NegNeg) were used as within-group factors, ‘group’ (patients vs. controls) as a between-group factor, and SUD value, VoC value, SAM valence value, SAM arousal value, M. zygomaticus magnitude, M. corrugator magnitude, SCL, and mean HR as dependent factors. For the physiological parameters, difference values between script 2 (negative, neutral, or positive) and script 1 (consistently negative) were calculated. For SCL and mimic muscle activity, a linear transformation [x – (Min + 0,1)] was conducted to avoid negative values.

For all analyses, p-values <0.05 (two-sided) were considered statistically significant. In cases in which Bonferroni corrections for multiple measurements were necessary, the calculated p-value was multiplied by the number of measurements to account for the increased statistical significance. In cases in which sphericity could not be assumed, the Greenhouse-Geisser correction for degrees of freedom was used. Post hoc tests were calculated as needed.

## RESULTS

### Intervention effect

The influence of both potential effect components, ‘valence switch type’ (NegPos vs. NegNeut vs. NegNeg) and ‘stimulation type’ (BLS vs. no BLS), was investigated by using a three-factorial ANOVA design with ‘group’ (patients vs. controls) as a between-factor. Only for physiological parameters, ‘phase’ (reading vs. imagining) was added as an additional within-factor. As the main effects for ‘phase’ were not significant (M. zygomaticus: [F(1,24) = 1.56, p = .224]; M. corrugator: [F(1,25) = 1.88, p = .183]; SCL: [F(1,34) = 3.38, p = .075]; HR: [F(1,24) = 1.20, p = .285]), all analyses were calculated by the averaged values.

### Subjective effects

#### SUD value

The findings are first reported for the SUD value (ranging from 0, indicating minimal distress, to 10, indicating maximal distress), which measures the current level of distress through the stressful event at the end of each intervention block (Fig. 3).

**Figure 3:**
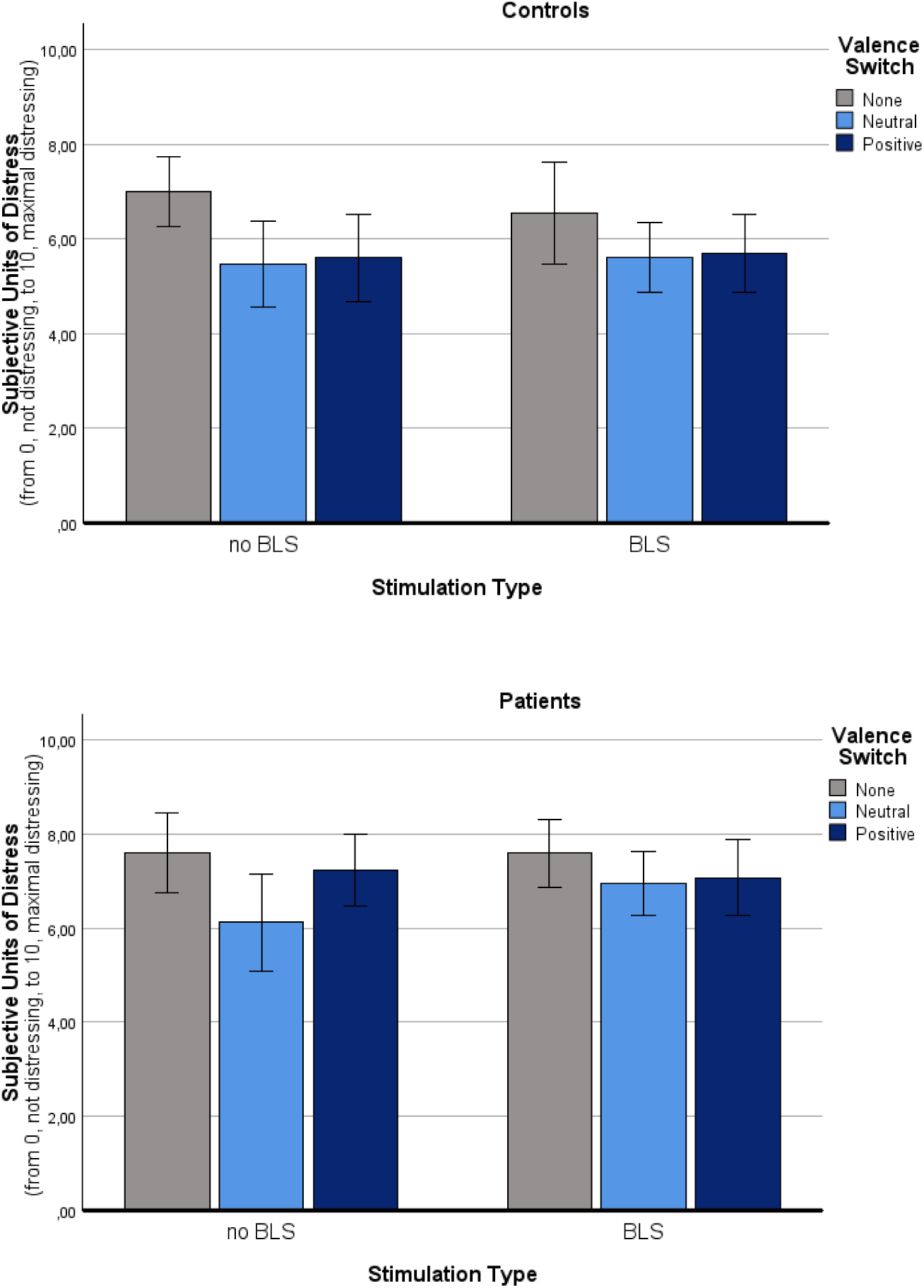
Intervention effects for the Subjective Units of Distress (SUD) score Changes in the Subjective Units of Distress (SUD) score for the two stimulation modes (BLS v. no BLS) and the three valence switch types (no valence shift vs. valence shift to neutral vs. valence shift to positive) in both groups (patients, controls). The SUD score is ranging from 0, minimal distress, to 10, maximal distress. Values are means with standard error bars.

***Main effect ‘group’.*** The factor ‘group’ significantly influenced the SUD rating of the participants [F(1,29) = 6.85, p = .014]. Post hoc tests indicated that the patients showed a significantly higher SUD value compared to the controls.

***Main effect ‘stimulation type’.*** The main effect for ‘stimulation type’ was not significant [F(1,29) = 0.53, p = .474], i.e., no difference was found between blocks with BLS and without BLS. No ‘stimulation type’ x ‘group’ interaction effect was found [F(1,29) = 2.09, p = .159].

***Main effect ‘valence switch type’.*** There was a significant main effect for ‘valence switch type’ [F(2,58) = 23.10, p < .001], with NegPos (p < .001) and NegNeut switches (p < .001) inducing a significant decrease of the SUD value compared to blocks without a valence change (NegNeg). There was a stronger SUD value decreasing effect of NegNeut switches compared to NegPos switches, but this difference did not fully reach the level of significance (p = .056). No ‘valence switch type’ x ‘group’ interaction was found [F(2,58) = 2.03, p = .140].

***‘Stimulation type’ x ‘group’ interaction effect.*** The interaction effects between ‘valence switch type’ x ‘stimulation type’ [F(2,58) = 1.12, p = .333] and ‘valence switch type’ x ‘stimulation type’ x ‘group’ [F(2,58) = 0.50, p = .608] were not significant.

#### VoC value

The findings are next reported for the VoC value (ranging from 1, minimal validity of the PC, to 7, maximal validity of the PC) at the end of each intervention block (Fig. 4).

**Figure 4:**
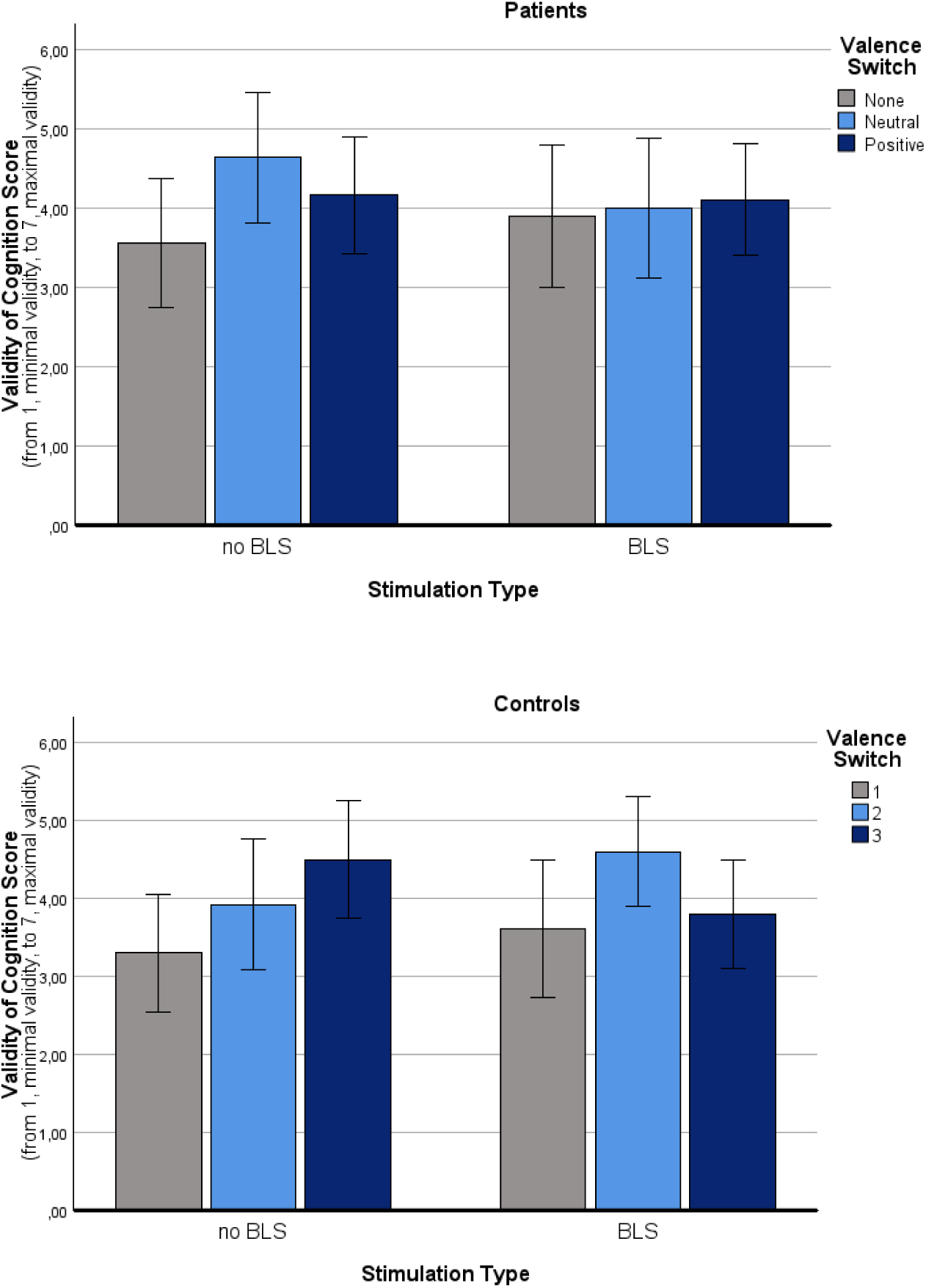
Intervention effects for the Validity of Cognition (VoC) score Changes in the Validity of Cognition (VoC) score for the three valence switch types (no valence shift vs. valence shift to neutral vs. valence shift to positive) and stimulation modes (BLS vs. no BLS) in both groups (patients, controls). Values are means with standard error bars.

***Main effect ‘group’:*** ‘Group’ did not influence the VoC rating findings [F(1,29) = 0.07, p = .794].

***Main effect ‘stimulation type’.*** The ‘stimulation type’ also did not influence the VoC ratings of the participants [F(1,29) = 0.01, p = .921], i.e., there was no difference between blocks with and without BLS. The interaction effect for ‘stimulation type’ x ‘group’ was not significant [F(1,29) = 1.20, p = .282].

***Main effect ‘valence switch type’.*** However, a significant main effect for the ‘valence switch type’ was found [F(2,58) =11.70, p < .001]. Post hoc analyses revealed that NegPos switches (p = .004) and NegNeut switches (p < .001) induced a significant increase in the VoC value compared to blocks without a valence change (NegNeg). No significant difference between NegPos and NegNeut scripts was found (p = .829), and there was no ‘valence switch type’ x ‘group’ interaction effect [F(2,58) = 0.465, p = .630].

***‘Stimulation type’ x ‘valence switch type’ interaction effect.*** No interaction effects were found, neither for ‘valence switch type’ x ‘stimulation type’ [F(1,58) = 1.13, p = .330] nor ‘valence switch type’ x ‘stimulation type’ x ‘group’ [F(1,58) = 2.24, p = .115].

#### SAM valence score

Intervention-based changes in subjective mood (Fig. 5) were measured via the SAM valence score (ranging from 1, unpleasant, to 9, pleasant).

**Figure 5:**
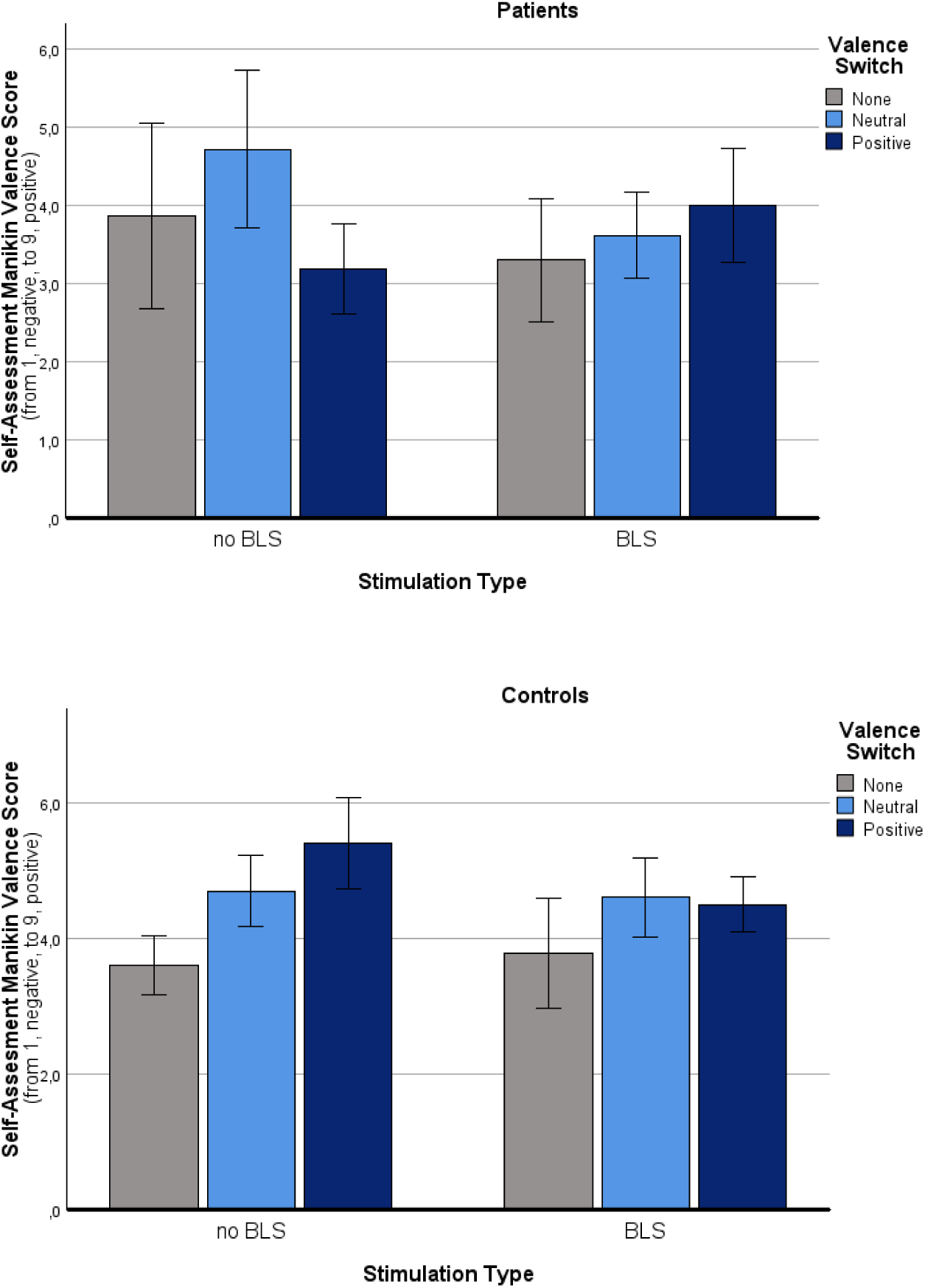
Intervention effects for the Self-Assessment Manikin (SAM) valence score Changes in the Self-Assessment Manikin (SAM) valence value for the three valence switch types (no valence shift vs. valence shift to neutral vs. valence shift to positive) and stimulation modes (BLS vs. no BLS) in both groups (patients, controls). The SAM valence scale is ranging from 1, negative to 9, positive. Values are means with standard error bars.

***Main effect ‘group’.*** The main effect of ‘group’ was significant [F(1,29) = 5.58, p = .025], with patients reporting lower SAM valence values, indicating a more negative mood.

***Main effect ‘stimulation type’.*** ‘Stimulation type’ did not influence the valence ratings of the participants [F(1,29) = 2.85, p = .102], i.e., there was no difference between script pairs with BLS and without BLS. The ‘stimulation type’ x ‘group’ interaction was not significant [F(1,29) = .01, p = .967].

***Main effect ‘valence switch type’.*** There was a significant main effect for ‘valence switch type’ [F(2,58) = 9.38, p < .001]. Post hoc analyses revealed that script pairs changing from negative to positive (NegPos, p < .001) and neutral (NegNeut, p = .006) were assessed significantly more positively (with a higher SAM valence value) than script pairs without a valence change. No difference was found between NegNeut and NegPos script pairs (p > .999). A significant ‘valence switch type’ x ‘group’ interaction effect was observed [F(2,58) = 5.71, p = .005]. In the control sample, the main effect for valence switch type remained significant [F(2,30) = 10.72, p < .001], whereas in the patient sample, only a statistical tendency was found [F(2,28) = 3.59, p = .082]. This interaction was particularly impressive in the NegPos condition: Here the patients showed significantly lower SAM valence values (indicating a more negative mood) than the controls did (p < .001). No group differences were found for script pairs with NegNeut changes (p = .582) or without a valence change (NegNeg, p > .999).

***‘Stimulation type’ x ‘valence switch type’ interaction effect.*** There was no significant ‘stimulation type’ x valence switch type’ interaction in the overall sample [F(2,58) = .76, p = .474] and also not separately for both groups (patients: [F(2,28) = 2.66, p = .176]; controls: [F(2,30) = 3.56, p = .082]), but a significant interaction effect between ‘stimulation type’ x ‘valence switch type’ x ‘group’ was found [F(2,58) = 5.14, p = .009]: In the patients, after application of BLS, the SUD value decrease (indicating negative mood) during the NegPos switches was no longer apparent. In the controls, under BLS, an unexpected decrease of the SAM valence value (indicating negative mood) was found.

#### SAM arousal score

Intervention-based changes in subjective arousal (Fig. 6) were measured via the SAM arousal scale (ranging from 1, low arousal, to 9, high arousal).

**Figure 6:**
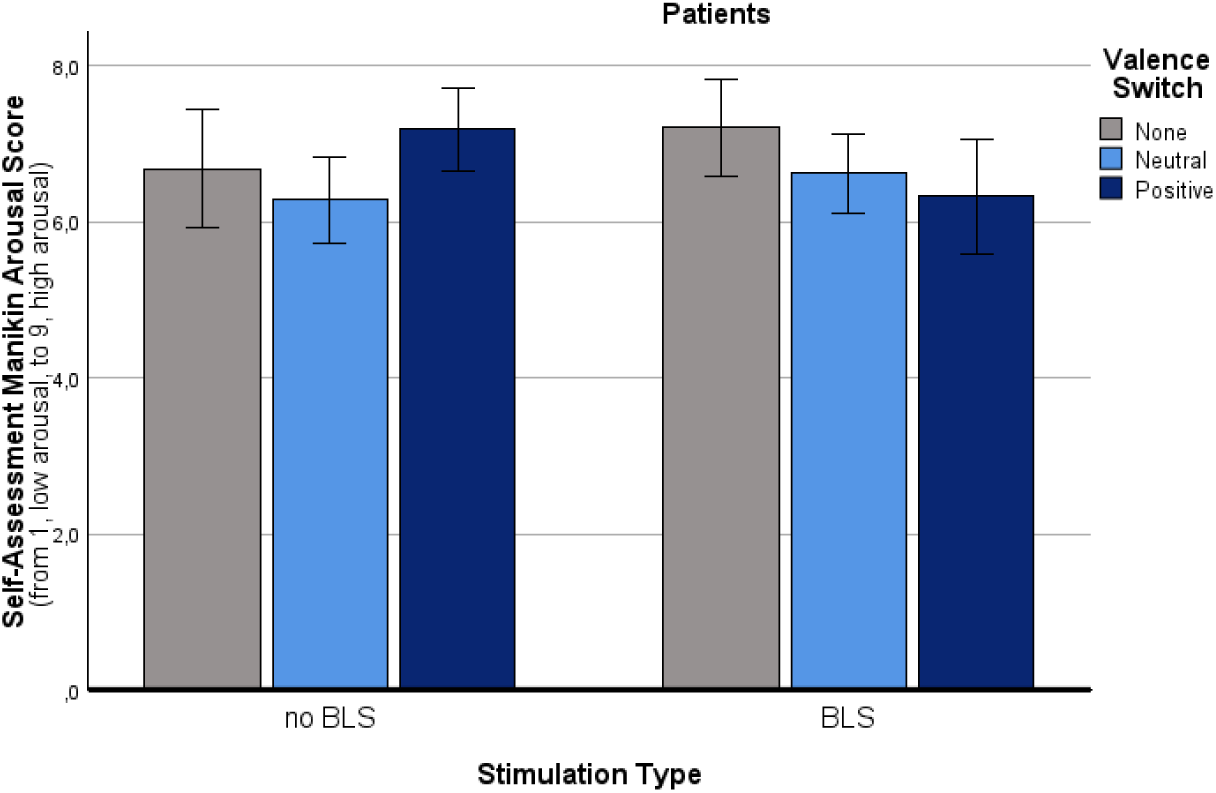
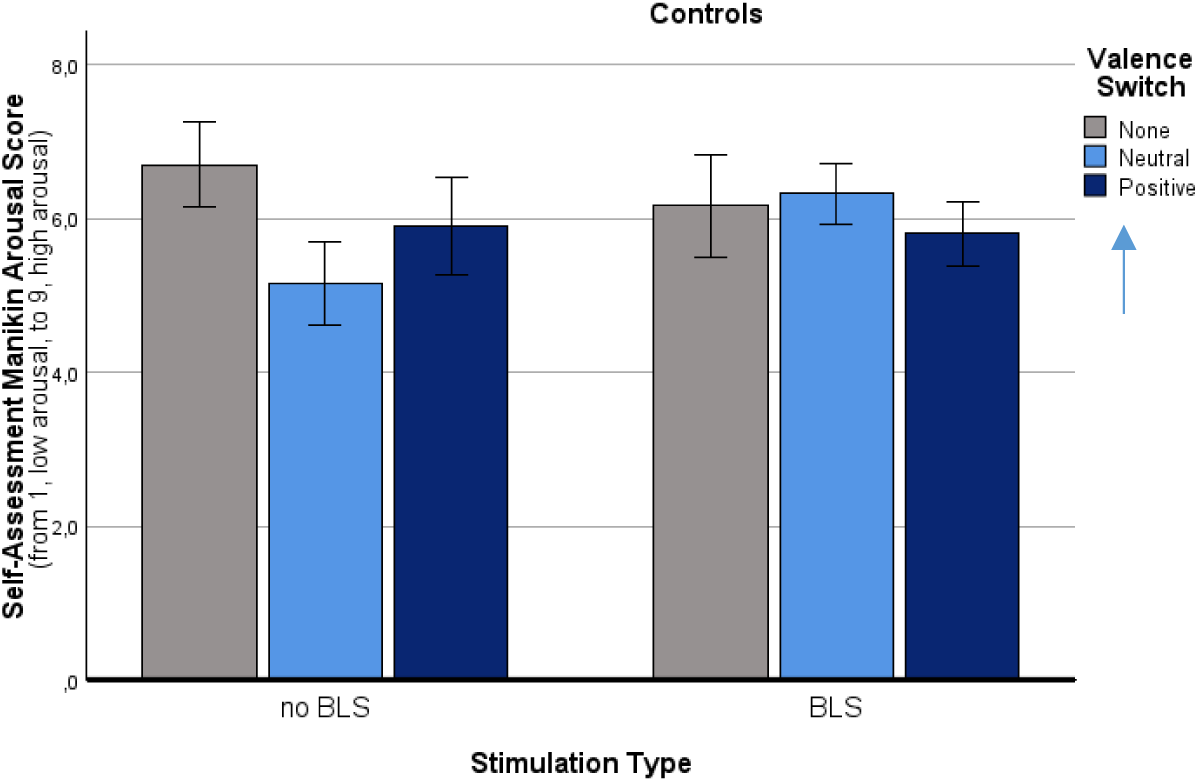
Intervention effects for the Self-Assessment Manikin (SAM) arousal score Changes in the Self-Assessment Manikin (SAM) arousal value for the three valence switch types (no valence shift vs. valence shift to neutral vs. valence shift to positive) and stimulation modes (BLS vs. no BLS) in both groups (patients, controls). The SAM arousal scale is ranging from 1, low arousal, to 9, high arousal. Values are means with standard error bars.

***Main effect ‘group’:*** ‘Group’ significantly influenced the SAM arousal rating of the participants [F(1,29) = 9.28, p = .005]. Post hoc tests indicated that the patients showed a significantly higher SAM arousal value (indicating a higher arousal) compared to the controls. ***Main effect ‘stimulation type’.*** ‘Stimulation type’ did not influence the subjective arousal ratings of the participants [F(1,29) = 0.76, p = .391], i.e., there was no difference between script pairs with BLS and without BLS. The ‘stimulation type’ x ‘group’ interaction [F(1,29) = 0.84, p = .391] was not significant.

***Main effect ‘valence switch type’.*** The valence switch type, in contrast, had a significant influence on the participants’ subjective arousal ratings [F(2,58) = 6.99, p = .002]. Script pairs changing into neutral (NegNeut) were assessed as significantly less arousal-provoking than script pairs without a valence change (NegNeg, p = .003). The same tendency was observed for NegPos scripts, although this effect did not reach significance (p = .065). No significant difference was found between the NegPos and NegNeut scripts (p = .606). No significant interaction between ‘valence switch type’ x ‘group’ was found [F(2,58) = 0.75, p = .475].

***‘Stimulation type’ x ‘valence switch type’ interaction effect.*** A significant ‘valence switch type’ x ‘stimulation type’ interaction effect was found [F(2,58) = 4.98, p = .010]: In the overall sample, an unexpected arousal-increasing effect of BLS compared to no BLS for script pairs changing into neutral was observed [F(1,29) = 16.36, p < .001]. This effect was significant for the control sample [F(2,30) = 7.56, p = .004], but did not appear in the patient group [F(2,28) = 2.67, p = .087], i.e. there was a significant interaction effect for ‘valence switch type’ x ‘stimulation type’ x ‘group’.[F(2,58) = 3.72, p = .030]: In the patients, an arousal-modulating effect for the NegPos scripts was observed: In the Non-BLS-condition, subjective arousal was paradoxically increased (p = .056, statistical tendency). Under BLS, this arousal increase was no longer apparent (p = .344), i,e. BLS diminished this paradoxical arousal-increasing effect. Additionally, there was a tendency for an arousal decrease for NegNeut scripts compared to NegNeg scripts in the patients (p = .084).

### Physiological effects

Intervention effects on physiological measures were calculated by subtracting the mean activity during script 1 from the mean activity during script 2.

#### M. zygomaticus activity

The M. zygomaticus activity is an emotional valence measure which indicates the extent of smiling. The findings are depicted in Fig. 7.

**Figure 7:**
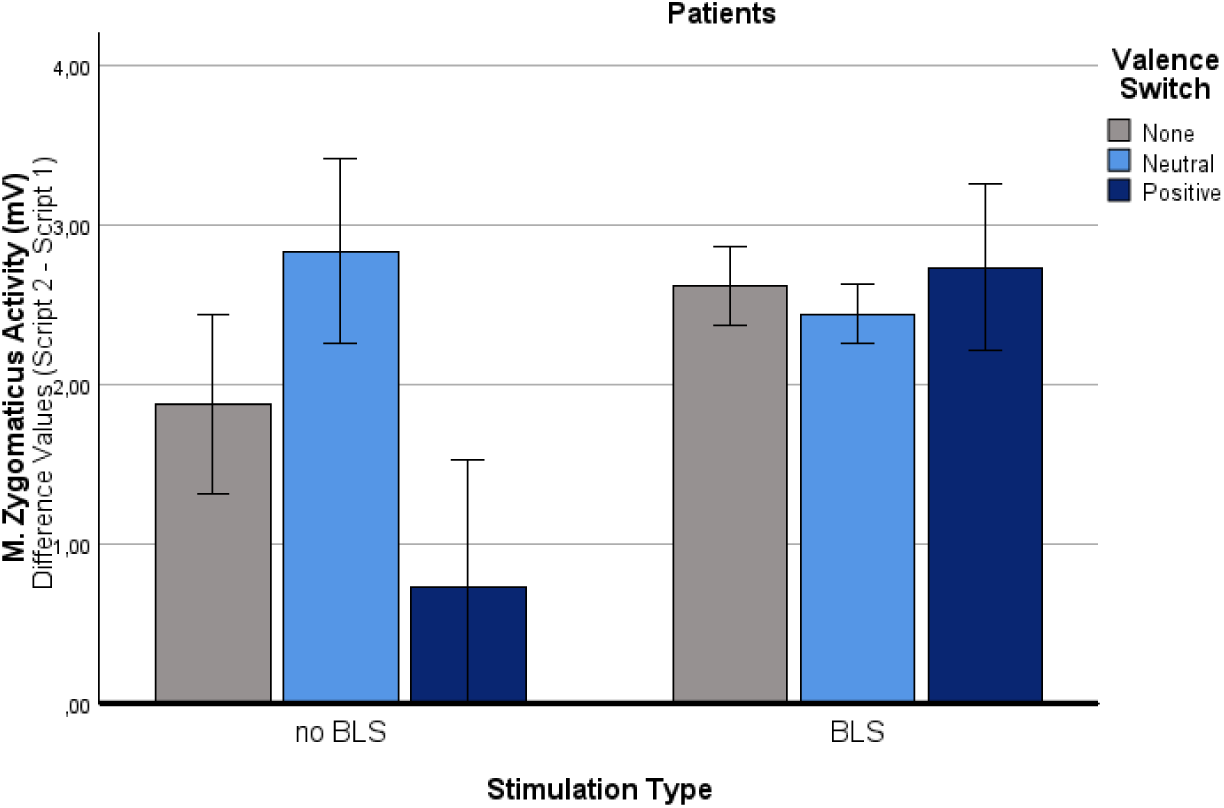
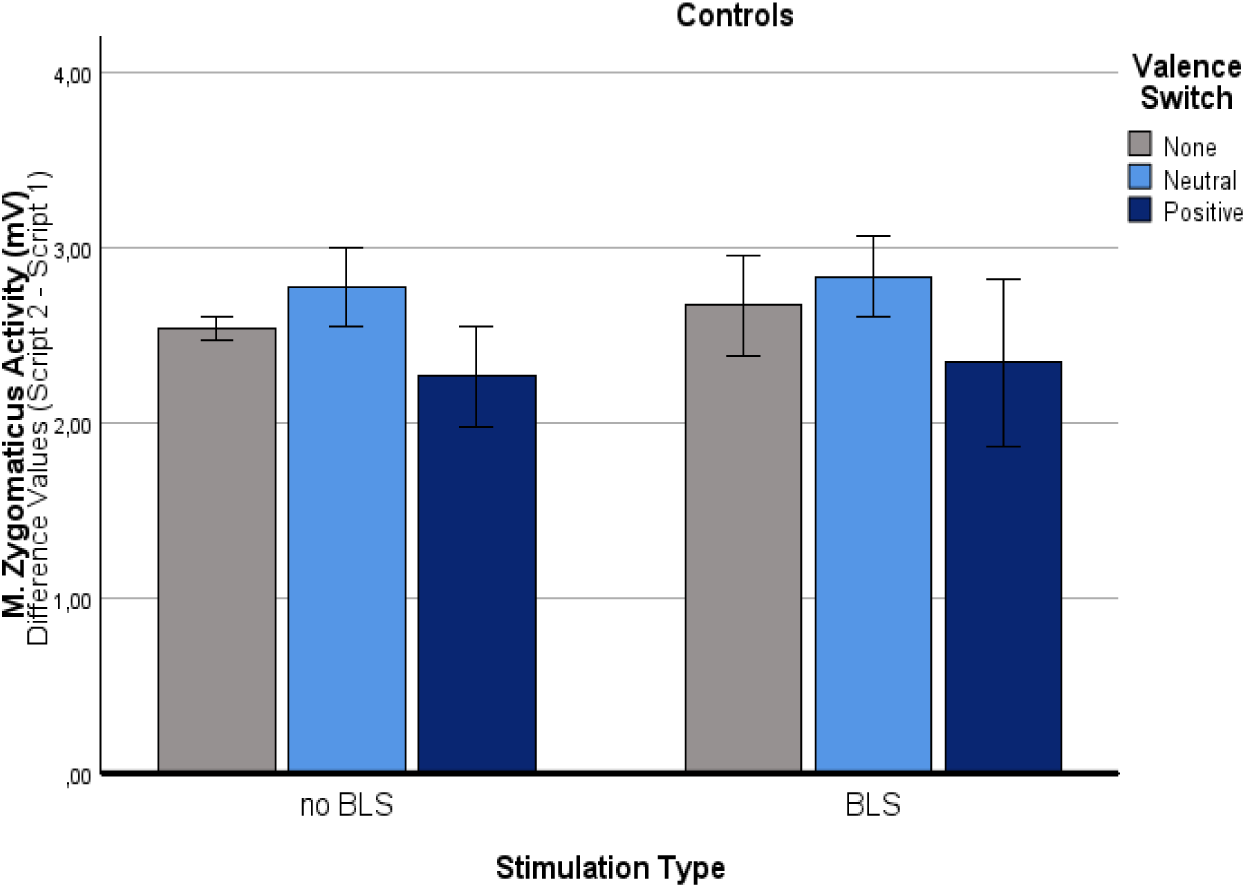
Interaction effects for the M. zygomaticus activity Changes in the M. zygomaticus activity for the three valence switch types (no valence shift vs. valence shift to neutral vs. valence shift to positive) and stimulation modes (BLS vs. no BLS) in both groups (patients, controls). Values are differences between the means of script 2 and 1 with standard error bars.

***Main effect ‘group’:*** ‘Group’ significantly influenced the M. zygomaticus activity of the participants [F(1,24) = 15.16, p < .001]. Post hoc tests revealed that the patients exhibited significantly lower M. zygomaticus activity (indicating less smiling) compared to the controls. ***Main effect ‘stimulation type’.*** A significant main effect for ‘stimulation type’ was found [F(1,24) = 18.68, p < .001]. Post hoc tests indicated that script pairs with BLS induced a significant increase in M. zygomaticus activity compared to script pairs without BLS. However, this main effect was only significant in the patient group: Here, the impact of the stimulation type on the M. zygomaticus activity was significant [F(1,10) = 17.56, p_c_ = .004], whereas for the controls it was not [F(1,10) = .73, p = .816]. Consequently, a significant ‘stimulation type’ x ‘group’ interaction effect was found [F(1,24) = 11.72, p = .002].

***Main effect ‘valence switch type’.*** Valence switch type significantly influenced the M. zygomaticus activity of the participants [F(2,48) = 12.10, p < .001]. Post hoc analyses revealed that script pairs changing into neutral (NegNeut) induced a significant increase in M. zygomaticus activity (p < .001) compared to script pairs without a valence change (NegNeg). For script pairs changing into positive (NegPos), in contrast, an unexpected decrease in zygomaticus activity was found compared to NegNeg scripts (p = .053) and NegNeut scripts (p < .001). This effect was particularly impressive in the patient group, but also appeared in the control group (even to a lesser extent). The ‘valence switch type’ x ‘group’ interaction effect did not reach the level of significance [F(2,48) = 1.03, p =.367].

***‘Stimulation type’ x ‘valence switch type’ interaction effect.*** Significant interaction effects for ‘valence switch type’ x ‘stimulation type’ [F(2,48) = 12.97, p < .001] as well as for ‘valence switch type’ x ‘stimulation type’ x ‘group’ were found [F(2,48) = 12.63, p < .001]. The findings are first reported for the *overall sample.* For valence changes into positive (NegPos) [F(1,24) = 20.46, p < .001] and also for script pairs without a valence change (NegNeg) [F(1,24) = 8.90, p = .018], a significant M. zygomaticus-increase under BLS compared to no BLS was found. In contrast, for valence changes into neutral (NegNeut), it was not [F(1,24) = 2.18, p = .459]. Separate analyses for both groups revealed, that this interaction did not occur in the control sample [F(2,28) = .05, p > .999], but was specific for the *patient* sample [F(2,20) = 14.40, p_c_ < .001]: During NegPos scripts [F(1,10) = 27.20, p < .001] and – as a tendency - during NegNeg scripts [F(1,10) = 5.83, p = .072], but not during NegNeut switches [F(1,10) = 2.40, p = .304], a significant increase of the M. zygomaticus activity was found under BLS compared to no BLS. In other words, under BLS, the unexpected decrease in the zygomaticus script for the NegPos patients was no longer apparent.

#### SCL

The SCL is an emotional arousal measure. The findings are depicted in Fig. 8.

**Figure 8:**
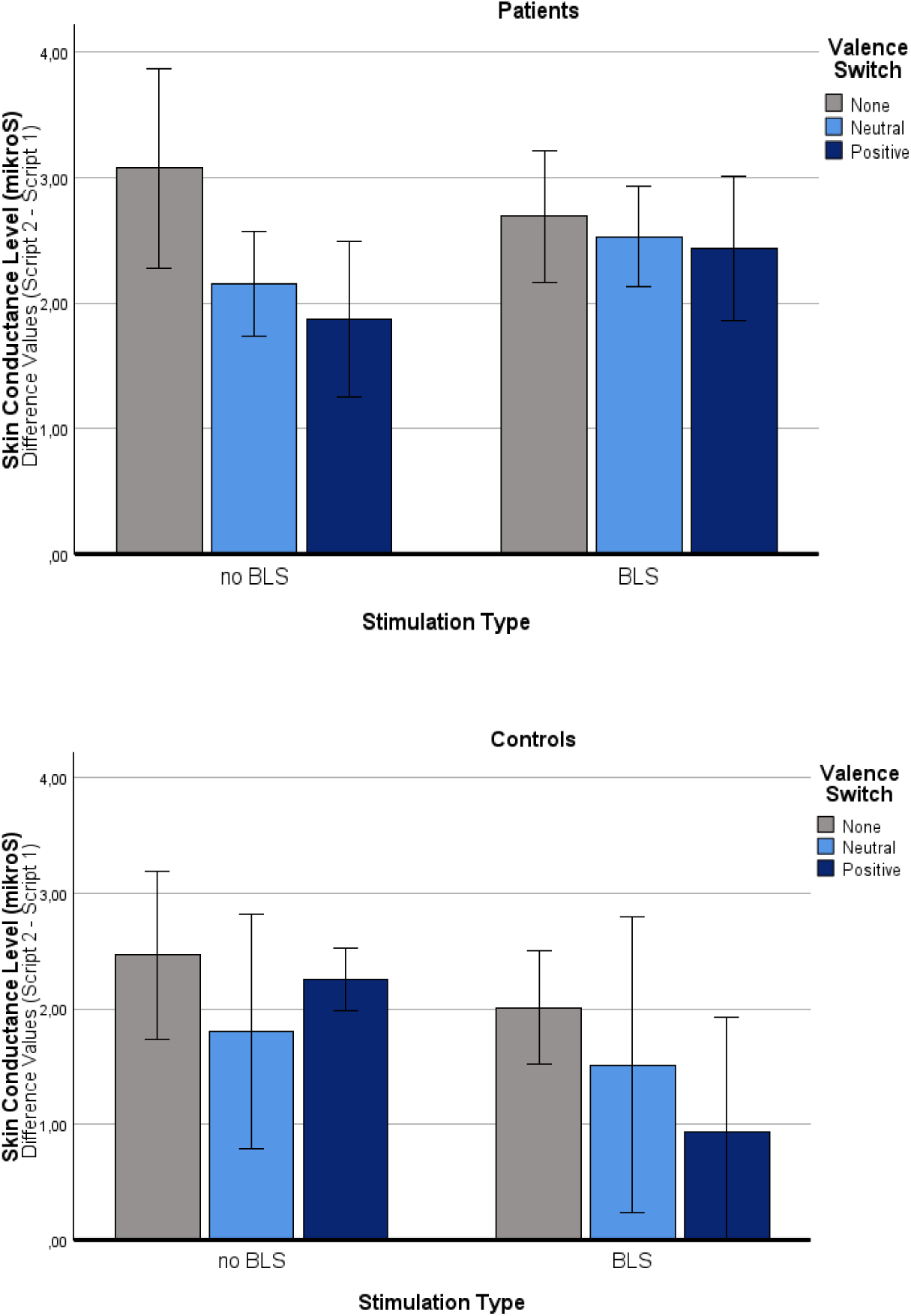
Intervention effects for the Skin Conductance Level (SCL) Changes in the skin conductance level for the three valence switch types (no valence shift vs. valence shift to neutral vs. valence shift to positive) and stimulation modes (BLS vs. no BLS) in both groups (patients, controls). Values are differences between the means of script 2 and 1 with standard error bars.

***Main effect ‘group’:*** ‘Group’ significantly influenced the SCL of the participants [F(1,34) = 5.56, p = .024]. Post hoc tests revealed that the patients exhibited a significantly greater SCL compared to the controls.

***Main effect ‘stimulation type’.*** In the overall sample, ‘stimulation type’ did not influence the SCL of the participants [F(1,34) = 1.84, p = .184], indicating no difference between script pairs with and without BLS. However, there was a significant interaction effect between ‘stimulation type’ x ‘group’: For the patients, the impact of the stimulation type on the SCL was not significant [F(1,16) = .60, p = .898]. For the controls, in contrast, there was a significant SCL-decreasing effect [F(1,18) = 6.04, p = .048], with BLS causing a substantial decrease compared to no BLS.

***Main effect ‘valence switch type’.*** The valence switch type had a significant influence on the SCL of the participants [F(2,68) = 3.86, p = .026]. Post hoc analyses revealed that valence changes into positive (NegPos) induced a significant decrease in SCL (p = .020) compared to scripts without valence changes, indicating a decreased arousal. In contrast, valence changes into neutral (NegNeut) did not (p = .244). No significant differences were found between NegNeut scripts and NegPos scripts (p > .999). There was no ‘valence switch type’ x ‘group’ interaction effect [F(2,68) = .03, p = .970].

***‘Stimulation type’ x ‘valence switch type’ interaction effect.*** The other interaction effects were not significant, neither for the ‘valence switch type’ x ‘stimulation type’ interaction [F(2,68) = .75, p = .478], nor for the ‘valence switch type’ x ‘stimulation type’ x ‘group’ interaction effect [F(2,68) = 2.46, p = .093].

#### M. corrugator activity

The M. corrugator activity is an emotional valence measure which indicates the extent of frowning. The findings are depicted in Fig. 9.

**Figure 9:**
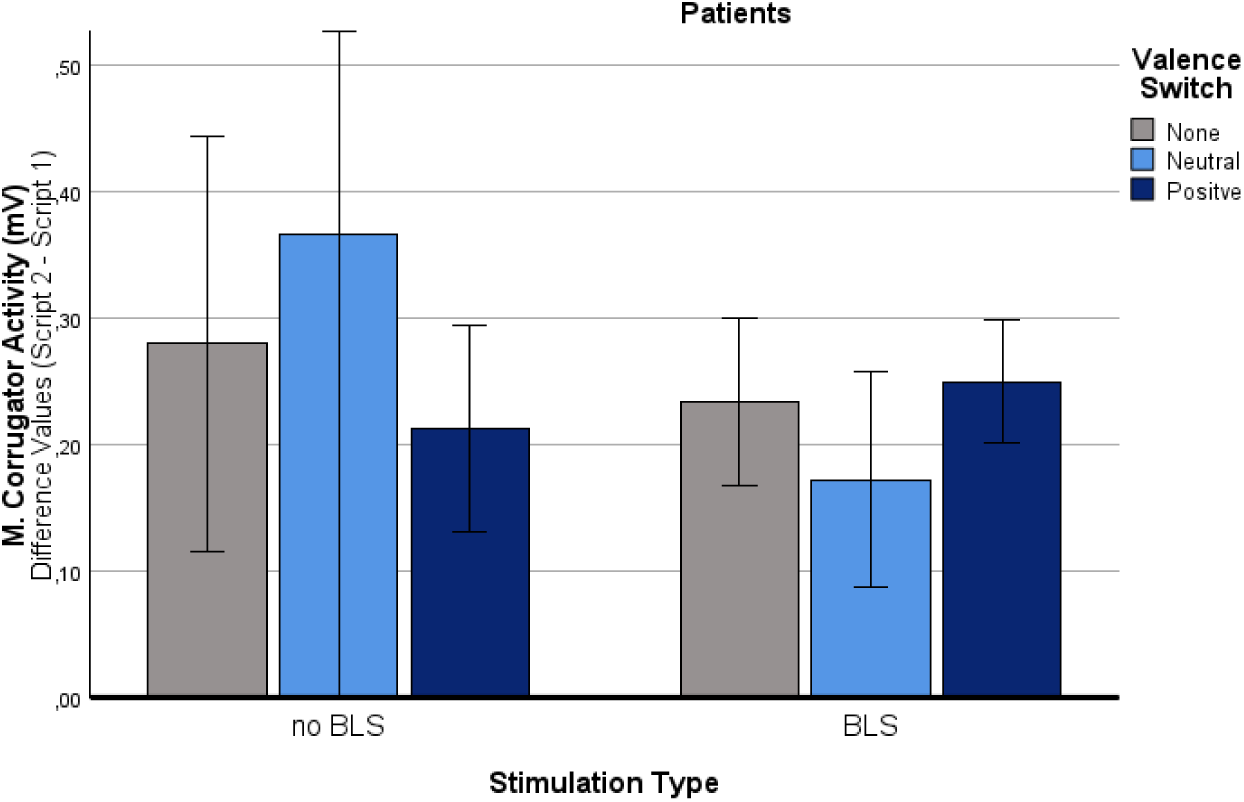
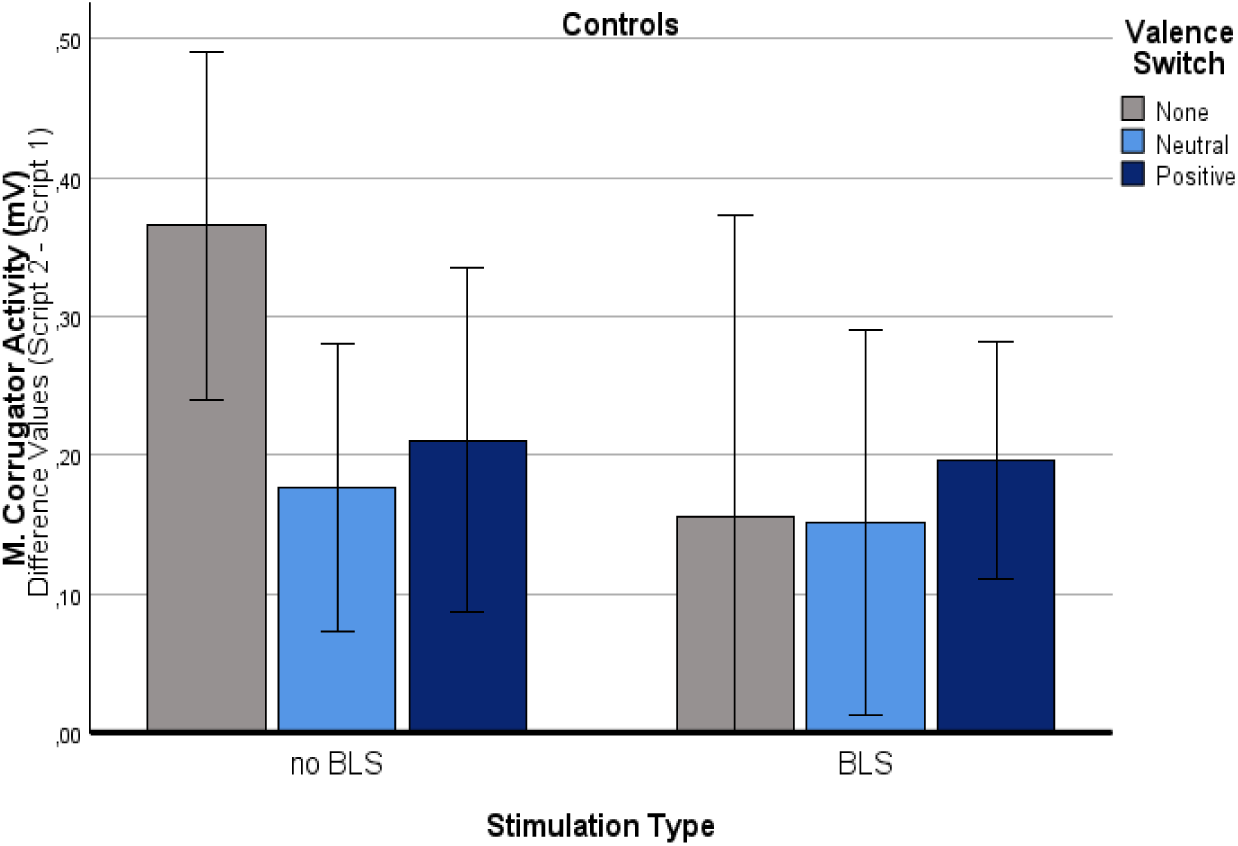
Intervention effects for the M. corrugator activity Changes in the M. corrugator activity for the three valence switch types (no valence shift vs. valence shift to neutral vs. valence shift to positive) and stimulation modes (BLS vs. no BLS) in both groups (patients, controls). Values are differences between the means of script 2 and 1 with standard error bars.

***Main effect ‘group’:*** ‘Group’ did not influence the M. corrugator activity of the participants [F(1,25) = .81, p = .377].

***Main effect ‘stimulation type’.*** There was a statistical tendency regarding the main effect for ‘stimulation type’ [F(1,25) = 3.18, p = .087]; script pairs with BLS induced a more substantial decrease in M. corrugator activity than script pairs without BLS. The ‘stimulation type’ x ‘group’ interaction effect was not significant [F(1,25) = .03, p = .860].

***Main effect ‘valence switch type’.*** The valence switch type did not significantly influence the M. corrugator activity of the participants [F(2,50) = 0.52, p = 0.533]. There was no significant interaction effect for ‘valence switch type’ x ‘group’ [F(2,50) = 0.70, p = .500].

***‘Stimulation type’ x ‘valence switch type’ interaction effect.*** The ‘valence switch type’ x ‘stimulation type’ [F(2,50) = .74, p = .481] and the ‘valence switch type’ x ‘stimulation type’ x ‘group’ interaction effect [F(2,50) = .93, p = .398] were not significant.

#### HR

The average HR can also provide some information about emotional valence: For example, a moderate increase (HR acceleration) is associated with positive emotions, and a decrease in (HR deceleration) is associated with negative emotions (Bradley, Codispoti, Cuthbert, & Lang, 2001). The findings are illustrated in Fig. 10.

**Figure 10:**
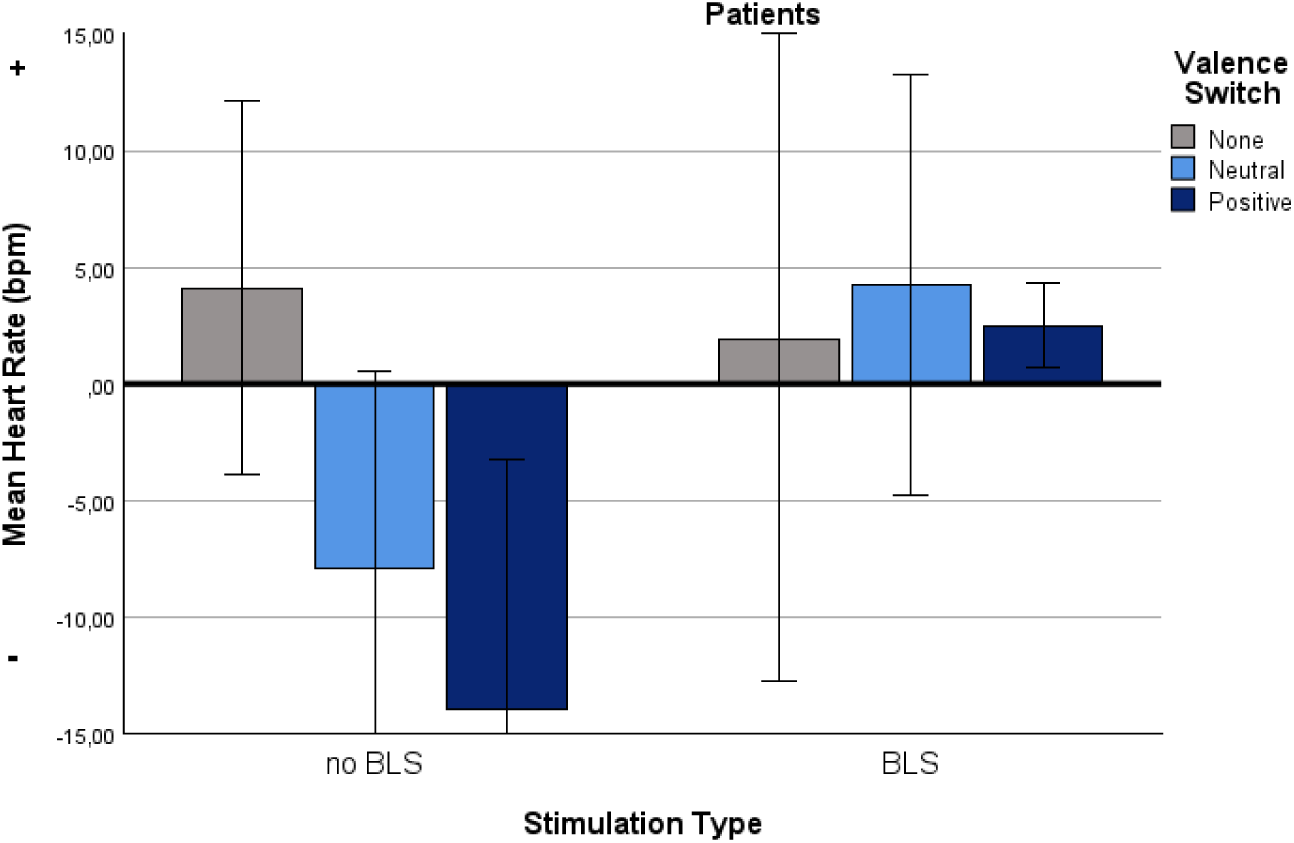
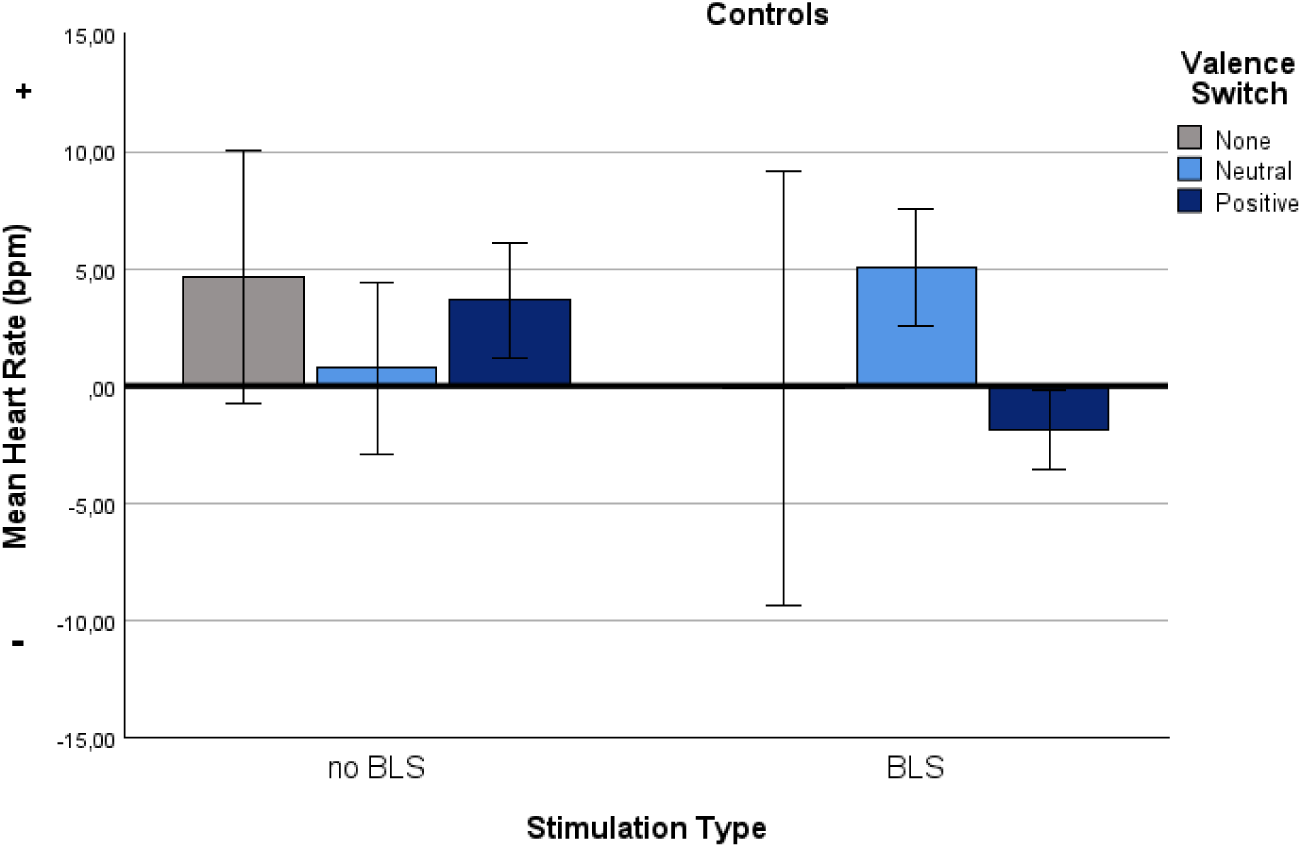
Intervention effects for the mean heart rate (HR) Changes in the mean heart rate (HR) for the three valence switch types (no valence shift vs. valence shift to neutral vs. valence shift to positive) and stimulation modes (BLS vs. no BLS) in both groups (patients, controls). Values are means and standard error bars.

***Main effect ‘group’:*** ‘Group’ did not influence the HR of the participants [F(1,24) = 1.97, p = .174].

***Main effect ‘stimulation type’.*** The main effect for ‘stimulation type’ [F(1,24) = 3.77, p = .064] showed a statistical tendency: A decrease in the HR for script pairs with BLS compared to those without BLS was found. In addition, there was a significant interaction effect between ‘stimulation type’ x ‘group’ [F(1,24) = 9.53, p =.005]: For the patients, the impact of the stimulation type on the HR was significant [F(1,10) = 9.33, p = .024], with an increased HR under BLS compared to no BLS. For the controls, no significant impact of the stimulation type on the HR was found [F(1,14) = .89, p = .726].

***Main effect ‘valence switch type’.*** The valence switch type had a significant influence on the HR of the participants [F(2,48) = 5.56, p = .018]. Post hoc analyses revealed that valence changes into positive (NegPos) induced a significant decrease in the mean HR compared to scripts without a valence change (NegNeg) (p = .036) and also compared to NegNeut script (p = .003). In contrast, no significant difference was found between NegNeut scripts and NegNeg scripts (p = .674). Additional analyses revealed an interaction effect between ‘valence switch type’ and ‘group’ (statistical tendency) [F(2,48) = 3.12, p = .065]: For the patients, the impact of the valence switch type on the HR was significant [F(2,20) = 5.08, p = .036]. For the controls, it was not [F(2,28) = .76, p = .428].

***‘Stimulation type’ x ‘valence switch type’ interaction effect.*** Neither the ‘valence switch type’ x ‘stimulation type’ interaction effect [F(2,48) = 3.12, p = .065] nor the ‘valence switch type’ x ‘stimulation type’ x ‘group’ interaction effect [F(2,48) = 2.08, p = .147] reached the level of significance, but separate analyses for the patients, corrected for multiple measurements, revealed differential effects. This effect was particularly pronounced during the NegPos switches, in which a significant BLS-induced increase of the HR was found (p = .028). Under BLS, the HR deceleration – as observed under no BLS - was no longer apparent, and, instead, a HR acceleration was observed. For NegNeut scripts, in contrast, the HR increase under BLS compared to no BLS did not reach the level of significance (p = .284). For NegNeg scripts, no significant influence of the stimulation type was observed (p > .999).

### Manipulation check

At the end of the simulated session, all single scripts were presented again in a random order - i.e., independently of their former pairing- and the participants were asked to subjectively rate them for emotional valence, arousal, and consistency as a manipulation check. A two-factorial ANOVA design with ‘script category’ (positive, neutral, negative) as a within-factor and ‘group’ (patients vs. controls) as a between-factor was used. The results showed significant changes in valence and arousal during confrontations with the different scripts in both groups. This indicates that the simulated valence changes subjectively induced real valence changes in the intended direction.

#### SAM valence score

For subjective mood (SAM valence score, ranging from 1, unpleasant, to 9, pleasant), a significant main effect for ‘script category’ [F(2,90) = 115.08, p < .001], no ‘script category’ x ‘group’ interaction effect [F(2,90) = 2.31, p = .105], and no main effect for ‘group’ was found [F(1,45) = .22, p = .639]. Positive scripts were estimated as significantly more enjoyable (p < .001) and negative scripts as significantly more distressing than neutral scripts (p < .001).

#### SAM arousal score

The analysis of the SAM arousal score also showed a significant main effect for ‘script category’ [F(2,90) = 32.34, p < .001], but no significant ‘script category’ x ‘group’ interaction effect [F(2,90) = .52, p = .565], and no main effect for ‘group’ [F(1,45) = 3.26, p = .078].

Negative scripts (p < .001) and positive scripts (p < .001) were rated significantly more arousal-provoking than neutral ones in both groups, and no significant difference between negative and positive scripts was found (p = .076).

#### SAM-analogue coherence Score

The analysis of the SAM-analogue coherence score showed a significant main effect for ‘script category’ [F(2,90) = 23.61, p < .001], no significant ‘script category’ x ‘group’ interaction effect [F(2,90) = .31, p = .705], and no main effect for ‘group’ [F(1,45) = .56, p = .458]: Negative scripts were experienced as more coherent as neutral scripts (p < .001), but did not differ significantly from positive scripts (p = .140). Positive scripts were experienced as more coherent than neutral scripts (p < .001).

## DISCUSSION

This pilot study compared PTSD patients and healthy volunteers with respect to their subjective and objective reaction patterns during a simulated EMDR intervention. Differential effects of valence switch type and stimulation mode were recorded in both groups for the first time.

### Basis group differences

Both groups differed significantly in terms of emotional arousal and valence during the experiment, with the patients reporting higher SUD values, higher arousal, and a more negative valence than the controls, indicating a higher event-related burden. In addition, they showed a lower M. zygomaticus activity (less smiling) and a higher SCL (higher arousal). This is in line with previous findings (McDonagh-Coyle et al., 2001) and with persistent hyperarousal as one criterion of PTSD (Sartory et al., 2013; Brewin, 2018).

### Influence of valence shift type and stimulation mood on the intervention outcome

#### Stimulation effect

Subjectively, there was *no* emotion-modulating effect of BLS. However, significant physiological effects were observed: In the patients, M. zygomaticus activity (i.e. smiling) was significantly increased under BLS compared to no BLS, and HR significantly accelerated, indicating an improvement in mood. These findings are in contrast to previous data demonstrating a physiological valence effect only in healthy individuals (Reichel et al., 2021) and mice (Jauch et al., 2023), but not in patients with PTSD (Pape et al., 2024). A possible explanation for these differences might be the use of autobiographical content in our study, whereas previous studies used standard scripts. In the controls, a BLS-induced decrease in SCL was observed. This finding aligns with previous studies (Elofsson et al., 2008; Montgomery et al., 1994; Sack et al., 2008; Schubert et al., 2010), which demonstrate physiological de-arousal during BLS. In both groups, a tendency for a decrease in M. corrugator activity (i.e. a reduction of frowning) under BLS was observed. Interestingly, all these processes seem to occur outside the patients’ conscious awareness.

#### Valence switch effect

The valence switch type significantly influenced the effectiveness of the intervention on both the physiological and subjective level: Subjectively, in blocks with a valence switch, a distress reduction in respect to the stressful event, an increased validity of the PC, a more positive mood, and a decreased arousal were found. In addition, significant physiological effects were observed, including an increase in smiling, a decrease in frowning, and a decreased SCL (de-arousal). Interestingly, the decrease in SCL was most substantial in script pairs simulating a valence switch into positive, whereas the M. zygomaticus effect was found only for valence switches into neutral. Each type of valence switch appears to have a distinct effect, creating a positive mood for the ‘negative to neutral’ type and reducing arousal for the ‘negative to positive’ type.

#### Interaction effects

Significant interaction effects were found between valence switch type, stimulation type, and group. This is first discussed for the *patients.* Here, in the Non-BLS condition, paradoxical effects were found for the NegPos switches, including an increase in subjective arousal and a worsening of subjective mood. This was also reflected on the physiological level, with the NegPos switches causing a decelerated HR and a decrease in M. zygomaticus activity, indicating a negative mood. I.e., despite its positive content, NegPos switches were subjectively experienced as *arousal-provoking* and *aversive* by the patients without BLS. There is already evidence from previous studies of a disturbed processing of positive stimuli in PTSD (Contractor et al., 2019; Litz, Orsillo, Kaloupek, and Weathers, 2000), and a persistent inability to experience positive emotion also belongs to the diagnosis criteria of PTSD following the DSM-5 (American Psychiatric Association, 2013). Our study provides first evidence that the application of BLS diminished these paradoxical effects, i.e., the patients were now able to perceive the NegPos switches as *positive* and *relieving*. In addition, a tendency for an improved processing (more smiling) of NegNeg scripts simulating ‘circling’ was found.

The *control*s, in contrast, did not require any BLS for experiencing NegPos and NegNeut switches as relieving and positive, i.e., they showed decreased arousal and an improvement in mood already in the no-BLS condition. BLS even had an irritating effect here. This was reflected in a subjective worsening of mood during NegPos switches and an increased subjective arousal during NegNeut switches in the BLS conditions compared to the no BLS condition. However, this can alternatively be interpreted as an increase in attention, in line with a previous study demonstrating a SCR-increasing effect of BLS during confrontation with positive standard scripts in healthy individuals (Reichel et al., 2021).

### Implications

From a *basic scientific perspective*, new insights into the mechanisms of action of the EMDR reprocessing phase in PTSD were created: The study replicated and expanded previous findings on objective BLS-induced physiological effects in patients with PTSD, and added novel findings by demonstrating valence switch effects and relevant interaction effects in the context of EMDR for the first time. The data shows that BLS triggers involuntary physiological processes that support a cognitive-affective re-appraisal of the traumatic event by a) increasing the positive effects of valence switches, b) allowing to switch from the trauma theme towards positive associations without a worsening of the mood, and c) activating involuntary positive feelings even in trials without valence switches. Interestingly, the participants were conscious of the valence switch effects under BLS, but not of the BLS effects per se. This might indicate, that the valence switch is necessary for the BLS effects to come into consciousness.

The findings also have *clinical implications*, for example by developing an AI- and biofeedback-supported tool that recognizes changes in valence based on physiological indicators during ongoing therapy sessions. Such a tool could be helpful to therapists, especially in bland cases in which it is not possible to detect the effect of BLS based on minimal nonverbal changes. A version for self-use by patients is also conceivable.

### Limitations

The study was conducted with a relatively small sample size, and some of the effects found in this sample were small. Larger sample sizes are required to utilize physiological data for clinical applications, such as those involving AI-based algorithms. Stimulation was performed via tactile vibration signals, and there is no information available on the interplay between BLS and valence switches for different stimulation types (such as visual stimulation). Even though autobiographical scripts were used instead of standard stimuli, and even though association pathways were simulated for the first time, the laboratory setting cannot be compared to a study in a clinical context. A human therapist was absent, and the association pathways were predetermined by the experimenter and not created by the patient themselves under BLS. Particularly regarding future clinical applications such as AI-supported biofeedback, further study seems essential to enable the real-time recording of physiological valence-change indicators during ongoing sessions. The selection of physiological indicators of valence changes is not exhaustive or complete. Other parameters, such as respiratory depth/rate or heart rate variability, also offer the potential to map BLS-induced valence change reactions and should be added in future studies. The time allotted for presenting the connections of each signaling pathway was very short. Many patients reported having too little time to grasp the content of the second script. The patients also seemed overwhelmed by the abrupt change, particularly with the NegPos scripts, which may have contributed to the adverse emotional reactions to this condition. For future simulated sessions, more time should be allocated for recording associations, and the transition should be smoother. Finally, the study aimed to examine the expanded mechanism of action of EMDR. However, this model could be further developed to include additional factors that may also influence the intervention’s success. Examples of this include the individual’s time and the number of associations required to reach the switching point under BLS, as well as the timely recognition of the switching point by the therapist and the patient’s subjective perception.

## Conclusion

This paper represents the first attempt to systematically measure the effects of various valence shift types and their interactions with different stimulation modes in research on EMDR. Regarding the presence or absence of BLS, previous findings on a physiological de-arousal in healthy individuals were replicated, and new findings were added, demonstrating an M. zygomaticus- and HR-acceleration-increasing effect of BLS in patients with PTSD for the first time. The results also show that the occurrence of valence shifts significantly influences the success of a simulated EMDR intervention, producing significant emotional changes ranging from de-arousal to a more positive mood, with both subjective and objective (i.e., physiological) parameters taken into account. Different types of valence shifts (i.e., changes from negative to positive vs. neutral) have differential effects, and a PTSD diagnosis significantly influences the findings. Significant interactions between valence switch type and stimulation mode were observed in the patients: Under BLS, valence switches into neutral had more substantial positive effects, and paradoxical adverse effects associated with sudden valence switches into positive were diminished. The findings demonstrate that valence shifts are relevant for the effect of EMDR as an additional effective factor and provide an empirical basis for AI-based clinical innovations, such as detecting valence shifts objectively via biofeedback.

## Author contributions

V.P. conceived and planned the experiment. O.W. supervised the project. C.K. and F.B. performed recruitment and data collection. C.F. contributed to the statistical data analysis. M.S., E.S., and O.W. contributed to the interpretation of the results. V.P. wrote the first draft of the manuscript, and all authors commented on previous versions of the manuscript. All authors read and approved the final manuscript. All authors have read and agreed to the published version of the manuscript.

## Data availability statement

The raw data of this article is freely available upon request.

## Ethics statement

The Ethics Committee of the University of Medicine Rostock approved the studies involving humans. The studies were conducted in accordance with the local legislation and institutional requirements. The participants provided their written informed consent to participate in this study.

